# Feasibility of satellite image and GIS sampling for population representative surveys: A case study from rural Guatemala

**DOI:** 10.1101/2020.08.20.20178632

**Authors:** Ann C. Miller, Peter Rohloff, Alexandre Blake, Eloin Dhaenens, Leah Shaw, Eva Tuiz, Francesco Grandesso, Carlos Mendoza Montano, Dana R Thomson

**Affiliations:** Department of Global Health and Social Medicine, Harvard Medical School; Boston MA USA; Division of Global Health Equity, Brigham and Women’s Hospital; Boston MA, USA; Wuqu’ Kawoq|Maya Health Alliance, Santiago Sacatepéquez, Guatemala; Epicentre, Paris, France; Institute of Nutrition of Central America and Panama (Instituto de Nutrición de Centroamérica y Panamá, INCAP), Guatemala City, Guatemala.; Department of Social Statistics and Demography, University of Southampton

## Abstract

**Background:** Population-representative household survey methods require up-to-date sampling frames and sample designs that minimize time and cost of fieldwork especially in low- and middle-income countries. Traditional methods such as multi-stage cluster sampling, random-walk, or spatial sampling can be cumbersome, costly or inaccurate, leading to well-known biases. However, a new tool, Epicentre’s Geo-Sampler program, allows simple random sampling of structures, which can eliminate some of these biases. We describe the study design process, experiences and lessons learned using Geo-Sampler for selection of a population representative sample for a kidney disease survey in two sites in Guatemala.

**Results:** We successfully used Epicentre’s Geo-sampler tool to sample 650 structures in two semi-urban Guatemalan communities. Overall, 82% of sampled structures were residential and could be approached for recruitment. Sample selection could be conducted by one person after 30 minutes of training. The process from sample selection to creating field maps took approximately 40 hours.

**Conclusion:** In combination with our design protocols, the Epicentre Geo-Sampler tool provided a feasible, rapid and lower-cost alternative to select a representative population sample for a prevalence survey in our semi-urban Guatemalan setting. The tool may work less well in settings with heavy arboreal cover or densely populated urban settings with multiple living units per structure. Similarly, while the method is an efficient step forward for including non-traditional living arrangements (people residing permanently or temporarily in businesses, religious institutions or other structures), it does not account for some of the most marginalized and vulnerable people in a population—the unhoused, street dwellers or people living in vehicles.

## Background

Population-representative household survey methods require up-to-date, accurate sampling frames and sample designs that minimize time and cost of fieldwork. These requirements are particularly important in low- and middle-income countries (LMICs) where other sources of data, such as census data and civil registration data are expensive to maintain, and likely to be out of date or incomplete(1). Routine national surveys including the Demographic Health Surveys (DHS) and Multiple Indicator Cluster Surveys (MICS) uniquely generate data needed for program and policy planning, monitoring development goals, and tracking development progress. However, they take place approximately every five years in most settings and may be out of date at time points of interest. Due to weak national data systems, household surveys are also often the main source of disease prevalence estimates in LMICs.

In 2017, while planning a household survey in two areas of Guatemala to estimate prevalence of chronic kidney disease of unknown etiology (CKDu), we considered available tools and methods for selecting a population-representative sample of households. At the time, the 2018 Guatemala Census was being planned, and only 2002 Census data were available. We therefore considered several additional options.

### Multi-stage cluster sampling

In multi-stage cluster samples, small areas are first selected at random from municipal or governmental maps, usually based on the last census, with probability proportionate to population size. The implied assumption is that the relative proportion of population in each small area – or “cluster” – has not changed substantially since the last census. In countries with extremely outdated census data, modelled gridded population estimates have instead been used to sample clusters(2). A complete enumeration of all households is then conducted in the selected clusters using foot travel and hand-drawn sketch mapping. Finally, households are randomly selected from within the enumerated clusters for inclusion in the study. While multi-stage cluster samples are widely considered to optimize statistical efficiency and fieldwork effort, sampling from either the available 2002 Guatemala census, or a gridded population dataset derived from this outdated census, were considered poor options.

### Multi-stage sampling with random-walk initiation

Other traditional methods, including the World Health Organization’s former Expanded Programme on Immunization (EPI) survey design, use two stage cluster samples(3). In the EPI design, the first stage clusters are selected with probability proportional to population size from census enumeration areas (government estimates); however, individual households are selected through a “spin-the-pen” and random walk mechanism. In this method, a pen or bottle is placed at a central location of the cluster and spun. Households in the indicated direction are identified, and one is selected at random to be the first household of the sample. The rest are selected in relation to that household in either a “next-nearest” fashion, or a skip pattern, in which, for example, every third or fifth house in the direction is sampled. This method, while commonly used for decades, can introduce bias(4, 5), as households nearest the centrally located start point will be more likely to be selected than households at a settlement’s periphery, and assumptions about non-response must be made; not every household has the same (or a known) probability of selection. This method was not suitable because it would require use of an outdated census simple frame and would not permit calculation of sample weights needed to produce unbiased estimates and confidence intervals of disease prevalence.

### Simple random sample of enumerated households

The most statistically efficient survey design would be a simple random sample of households from the entire population if an updated, complete household sample frame were available. Surveys with smaller coverage areas have selected simple random samples of households, for example in China, using electronic listings of households in two districts (6), and Democratic Republic of Congo (7) using civil registry data for household listings of small catchment areas. However, resource-limited countries like Guatemala rarely have a unique home address system or database of household GPS locations. Alternatively, satellite data can be employed to digitize all structures within an area (8), though this is time and labor intensive, and requires geographic skills and knowledge that are not always available in resource-poor settings.

### Spatial Samples

Other surveys have attempted to employ spatial sampling approaches, for example in setting up a cholera vaccination campaign in Democratic Republic of Congo (9), a basic pediatric health indicators survey in Zambia(10), or a diabetes prevalence survey in Guatemala(11). Often, these samples are constructed through generation of a regular geometric grid, from which points are selected through simple, systematic, stratified or clustered random sampling for inclusion in the study. Spatial samples have also been used to map risk to human health across space (e.g. pollution(12) or distribution of species (13)), and were combined with methods such as capture-recapture and adjustment for population density (13, 14).

Importantly, simple spatial samples do not result in population-representative samples because human populations are not distributed uniformly across geographic areas. Simple spatial sampling designs lead to oversampling of sparsely distributed households. In rural areas, this means that remote households will be more likely sampled than in settlements, and in urban areas, wealthier households, which often have larger areal footprints, will be more likely to be sampled than small, densely packed poor households, resulting in biased population samples and biased results. Stratification by population density prior to random spatial sampling has been used in population surveys(12), requiring population proportional to size sampling and weighting to account for that in analysis.

Epicentre, a Non-Governmental Organization working with Doctors Without Borders, has recently created a new sampling tool, known as Geo-Sampler, to assist researchers in generating household samples using satellite imagery, eliminating the need for cluster-level sampling. Geo-Sampler does this via a Google Earth-based interface by generating random points within a polygon (e.g. district, or city) superimposed on up-to-date satellite imagery. Users can optionally set a radius around the point (e.g. 10 meters to cover the size of a typical building), and incrementally select an infinite number of random points. Importantly, in the Geo-Sampler protocol, all points that do not include a structure are discarded, resulting in a sample of structures rather than geographic units, which overcomes the limitations of simple random spatial sampling (illustrated in Figure 1). Additional data about structure occupancy and population density are collected during the survey and used to generate sample weights that adjust for non-populated structures or structures with multiple households.

**Figure 1:**
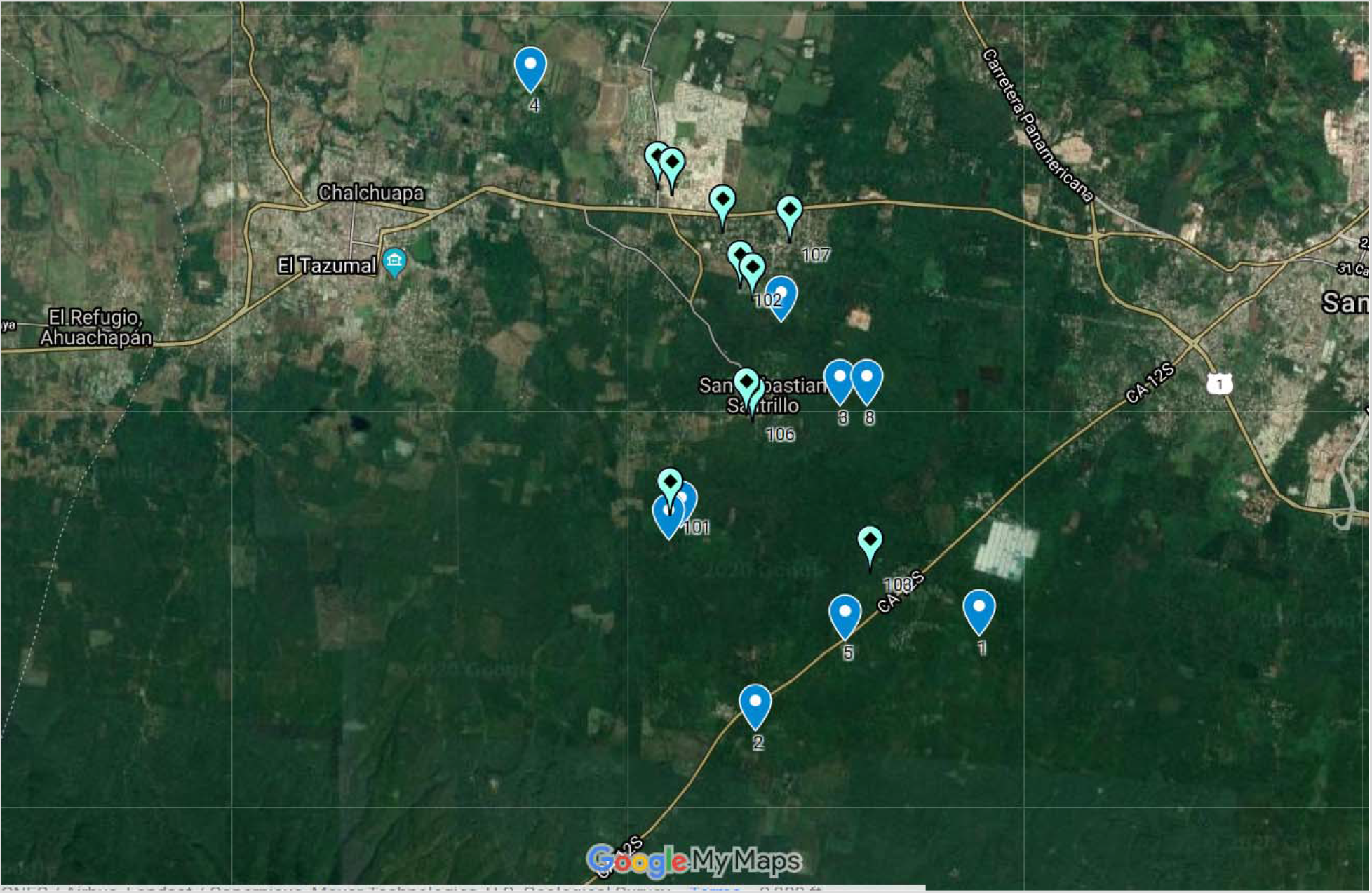
Example of Comparison of Simple Spatial Sampling (dark blue markers 1–10) vs. Simple random sample of structures (light blue markers 101–110) in which only those selected points that contain a structure were retained. Both were generated by Geo-Sampler.

To generate a population-representative sample from an up-to-date sample frame with maximum statistical power and efficient field protocols, we used Geo-Sampler with protocols adapted to our setting and study needs. Although some studies led by Epicentre have used earlier versions of Geo-Sampler to select households within a two-stage cluster design(15, 16), as yet, no articles detail its use as a tool to generate a full sample. Therefore, here we describe our experience with this new tool in designing and conducting a population survey for estimating the prevalence of chronic kidney disease in two areas of Guatemala.

## Methods

### Description of the main study and study area

This research is part of an National Institutes of Health R21-funded study (1R21TW010831–01) on chronic kidney disease of unknown origin (CKDu), which emerged as a recent epidemic in Central America and other global sites in individuals without traditional CKD risk profiles (young, male, without diabetes or hypertension)(17). The main objective of the study is to estimate the prevalence of both CKD and CKDu in two communities with different risk profiles. The study uses the infrastructure of the healthcare organization Wuqu’ Kawoq|Maya Health Alliance which has a long-standing presence in both study communities. Tecpán, Department of Chimaltenango (core population, excluding outlying settlements, approximately 84,000) is a majority indigenous Maya community in the temperate highlands of central Guatemala (elevation 7500 feet). San Antonio Suchitepéquez, Department of Suchitepéquez (core population approximately 52,000) is a lowland Pacific coastal warm-climate town with larger non-indigenous population (elevation 1000–1500 feet). The sites were chosen to provide a diversity of CKD risk factors, including differential exposure to heat stress and strenuous agricultural labor, profiles of pesticides in common use, diet diversity, risk for obesity and cardiovascular disease, and early life insults such as child malnutrition.

The prevalence survey was designed to be conducted through home visits by study nurses, consisting of an interview with any eligible household members (non-pregnant adults who agree to participate and provide informed consent), biometric measurements using a bioimpedance scale, urine samples to measure urine protein and creatinine, and serum samples to measure serum creatinine and glycosylated hemoglobin A1C. These test procedures allow for screening for diabetes and CKD and permit staging of any diagnosed CKD using Kidney Disease: Improving Global Outcomes(KDIGO) guidelines(18). Positive results are returned to the participants by the study team along with confirmatory testing and a facilitated referral to public health or specialty clinics, as needed, for any follow-up care.

### Constructing the sampling frame and random sampling of structures and households

An *a priori* sample size calculation of 700 people from 350 households was required to estimate the point prevalence of CKD with a margin of error of 0.35—220 households from Tecpán and 130 from San Antonio Suchitepéquez—including inflation for refusals, household clustering in structures, and an expected prevalence of 10%(19).

Using ArcGIS 10.15.1, we created polygon shapefiles for Tecpán and San Antonio Suchitepéquez. Shapefiles were drawn to include the municipal boundaries of each town, with some expansion at the edges to include households when town boundaries cut through a tight group of structures. These maps were reviewed and approved by the co-investigators with knowledge of the area (PR, CMM). We then imported the shapefiles into Geo-Sampler.

The Geo-Sampler tool (version 0.1.0.47 (2018–05–02)) interfaces with Google Earth and randomly selects points within a specified polygon shape, following a sampling with replacement technique. We set a buffer of 15m around each sampling point, approximately the size of a lot in these regions of Guatemala, to reduce the number of points that needed to be selected and potentially dropped. Sample selection was performed beginning in April 2018 for Tecpán and May 2018 for San Antonio Suchitepéquez, and the imagery for both study sites in Google Earth was dated January 9, 2018.

One of the co-authors (ACM) selected and reviewed each point (with 15m buffer) according to the following rules:

- If a structure existed within the buffer, the point was kept, and the structure within the buffer was included in the sample.
- If more than one structure existed within the buffer and the centroid point fell between them, the structure with any wall or corner nearest to the centroid point (based on visual estimation) was selected.

- If the closest structures were equidistant to the point (based on visual estimation), we selected the structure closest to 12:00, considering all structures in a clockwise fashion from 12:00.
- All sample structures were kept, regardless if they appeared to be non-residential, multi-family, or part of compounds (e.g., outdoor kitchen or privy).
- Additional points with 15m buffers were selected until the target sample size was reached.

By using a buffer around points, points did not always fall directly over a structure. To record sampled structure locations, we used Google Earth (same 1/9/2018 imagery) to manually digitize latitude and longitude coordinates for the sampled structures, and recorded these in a .xls spreadsheet file. Given potential structure abandonment and nonresidential structures, an extra 155 replacement structures were selected in Tecpán and 145 replacement clusters were selected in San Antonio Suchitepéquez. At the end of each Geo-Sampling session, a .csv file of retained sample points, and .xls file of corresponding sampled structures latitude and longitude coordinates were saved. Because sampling with replacement was used, each selected structure was compared with its nearest neighbors to be sure that the same structure was not included more than once.

Hard copy maps of each structure with a Google Earth base layer at a finer resolution were also provided to the data collectors for navigation.

### Approaching household and calculating sampling weights

The protocol for approaching households was as follows: the data collectors were given a list of selected structures with coordinates and maps from the main sample (n = 220 in Tecpán, and n = 130 in Suchitepéquez) and separate lists of coordinates and maps of replacement structures. If the structure was a residence, the data collectors initiated enrollment activities for a household. If the structure was not part of a residence, the data collectors attempted to identify the nearest residence, turning in a clockwise circle on the street in front of the structure. If none of the structures in that circle were residences (or if there were no other structures), the data collectors made a note and a structure from the replacement list was visited instead. If the structure was a residence that contained more than one household, a list of the households was made and one household was selected at random. For each household, queries outlined in Table 3 were collected in order to appropriately weight the survey results.

Sample weights were calculated as follows, using stratum (city), structure and household response rates. Notation below uses k for strata, j for structures and i for households.

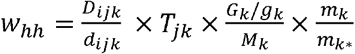

Where:

G_k_is the total population in stratum k (using estimated projections by the Guatemala National Institute of Statistics for the municipalities of Tecpán and San Antonio Suchitepéquez(20))

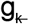is the average household size in stratum k as estimated by Guatemala DHS 2014–2015(21)

M_k_ is the number of target households in stratum *k*

D_ijk_ is the number of households *i* enumerated in structure *j* in stratum *k*

d_ijk_ is the number of households *i* selected in structure *j* in stratum *k*

T_jk_ is the number of structures *j* in household *i* in stratum *k*

m_k_ is the number of approached households in stratum *k* during fieldwork

m_k*_ is the number of responded households in stratum *k* during fieldwork

### Pilot Exercise

Prior to the initiation of formal data collection, a pilot was conducted with study data collectors to determine whether use of hand-held devices using readily available commercial GPS software (Google Maps) was feasible for data collectors following a brief training to identify households using latitude and longitude coordinates. Other methods were also tested, including the use of a Garmin GPS device and printed copies of maps. The training took one afternoon and consisted of didactics and examples. The pilot consisted of field work with the 2 data collectors and the study coordinator, who were given 10 structure coordinates to identify, and printed maps with the structures marked. Using their office-issued mobile Android phones (models Motorola MotoE4 Plus, Samsung Galaxy J3, Samsung A10) the nurses programmed the coordinates into the phones and used the factory-installed directional navigation software to find the structures, with real-time field assistance and as-needed support from one of the study investigators (PR). In this pilot, the study staff were able to program coordinates into the phones and easily find assigned structures, and so we elected to proceed with hand-held devices with Google Maps rather than other methods. This was cost-saving, as it was not necessary to purchase GPS devices. All staff were also already familiar with the use of Google Maps.

## Results

### Sample selection

Of the main sample of 350 structures, 288 (82%) were residential, 81 (23%) had more than one building per residence, and 17 (5%) had more than one family per structure (Table 1). This was different between the two sites. Tecpán reported statistically significantly more non-residential buildings (44, 20.0%) than San Antonio Suchitepéquez (15, 11.5%) in the main sample, and significantly more structures composing a residence (mean of 1.84 in Tecpán vs. 1.40 in San Antonio Suchitepéquez, p< 0.01). No differences were seen between the two sites with respect to the number of multi-household structures.

**Table 1:**
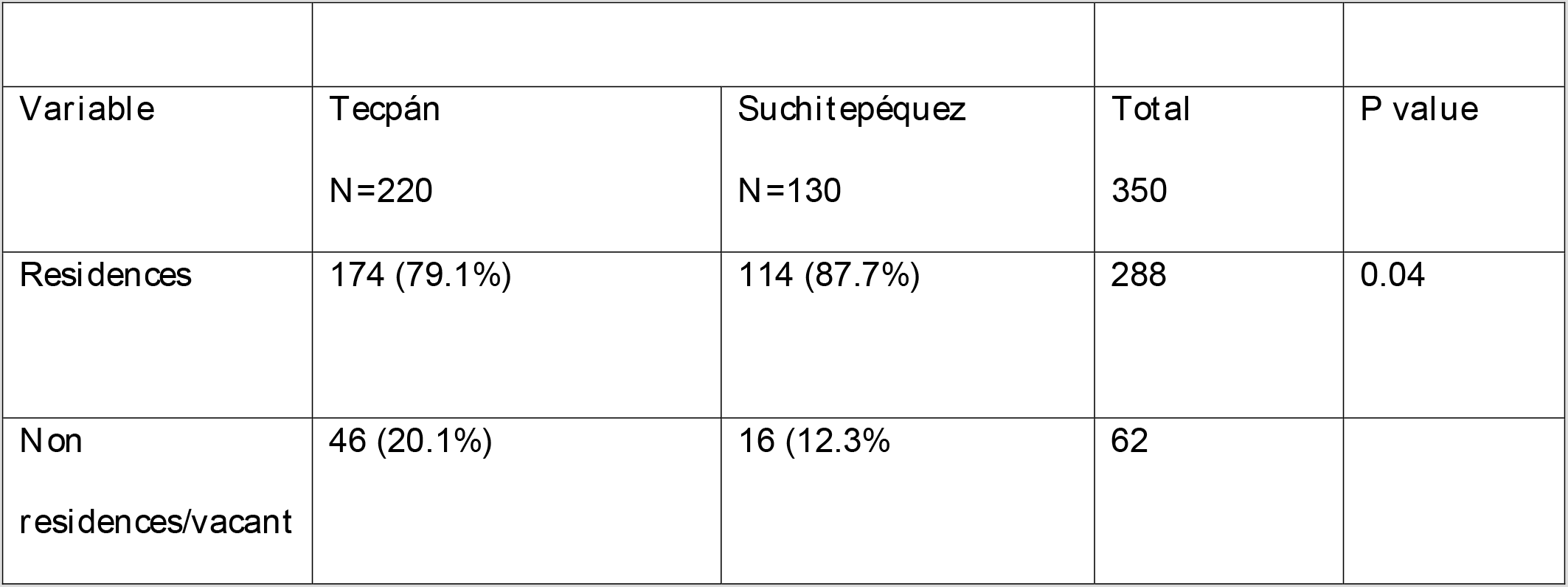
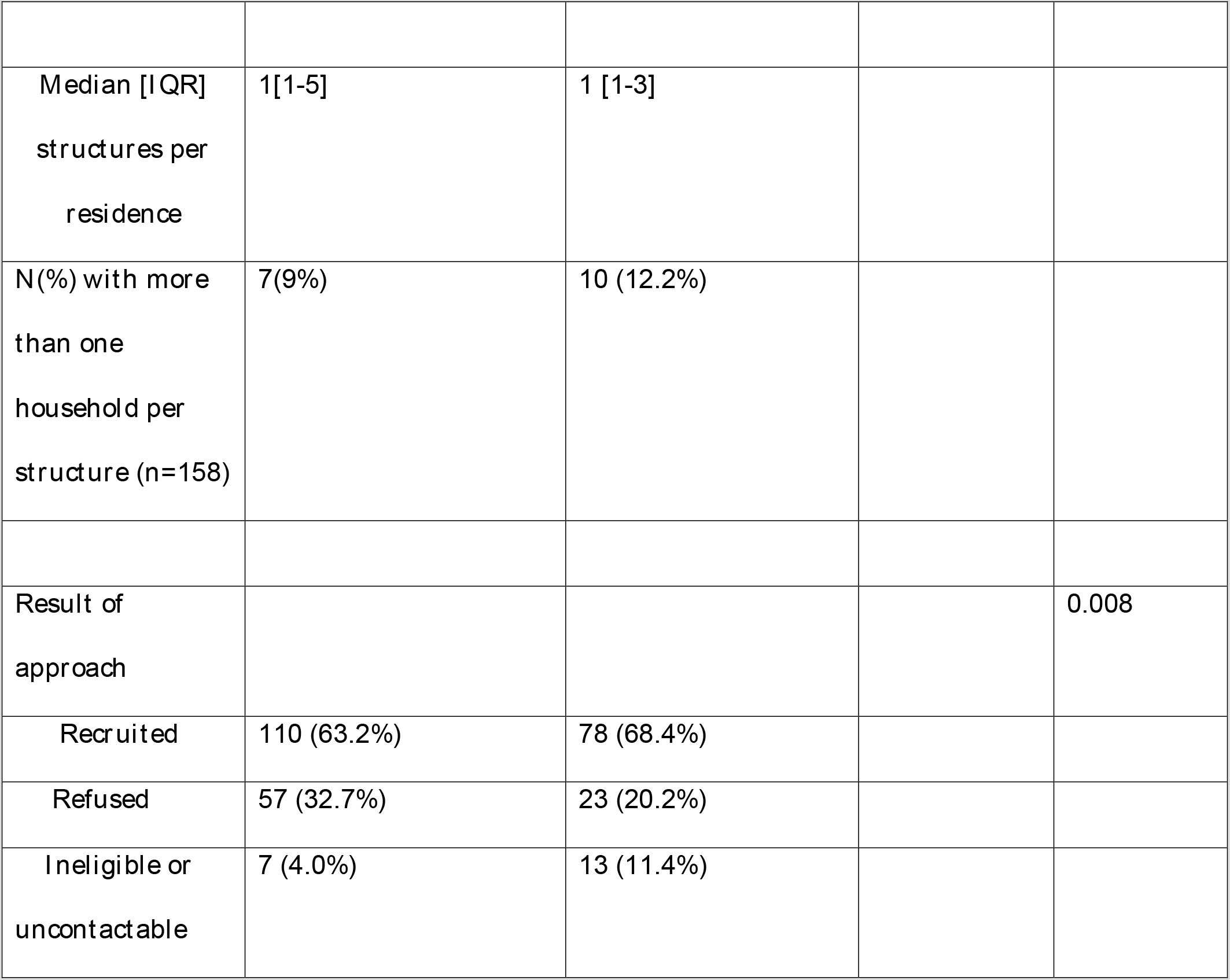
Results of approaches to households in CKD prevalence survey, Guatemala.

| Variable | Tecpán<br>N=220 | Suchitepéquez<br>N=130 | Total<br>350 | P value |
| --- | --- | --- | --- | --- |
| Residences | 174 (79.1%) | 114 (87.7%) | 288 | 0.04 |
| Non<br>residences/vacant | 46 (20.1%) | 16 (12.3%) | 62 |  |
| **Median [IQR]**<br>**structures per**<br>**residence** | **1[1-5]** | **1 [1-3]** |  |  |
| **N(%) with more**<br>**than one**<br>**household per**<br>**structure (n=158)** | **7(9%)** | **10 (12.2%)** |  |  |
| **Result of**<br>**approach** |  |  |  | **0.008** |
| **Recruited** | **110 (63.2%)** | **78 (68.4%)** |  |  |
| **Refused** | **57 (32.7%)** | **23 (20.2%)** |  |  |
| **Ineligible or**<br>**uncontactable** | **7 (4.0%)** | **13 (11.4%)** |  |  |

### Feasibility

We found sample selection using Epicentre’s Geo-Sampler tool to be feasible and practicable in this setting. Sample selection was conducted by one person (ACM), after receiving approximately 30 minutes of training on the Geo-Sampler tool from a contact at Epicentre (AB). Creating the sample dataset in our study required three steps (described in detail in the Methods section): selecting the point-with-buffer sample, recording sampled structure coordinates, and creating field maps, although updates to Geo-Sampler now allow the structure coordinates to be created and downloaded to a spreadsheet within the tool. The process of selecting the 650 main and replacement coordinates took approximately 24 hours. The process of manually digitizing and recording sampled structure coordinates took approximately another 12.5 hours. The process of creating field maps took about two hours, for a total of 38.5 hours. These skills required for this work were limited, and can be performed by anyone with basic spreadsheet skills, familiarity with Google Earth, an initial training in the Geo-Sampler tool, and sufficient familiarity with geographic coordinate systems.

**Table 2:**
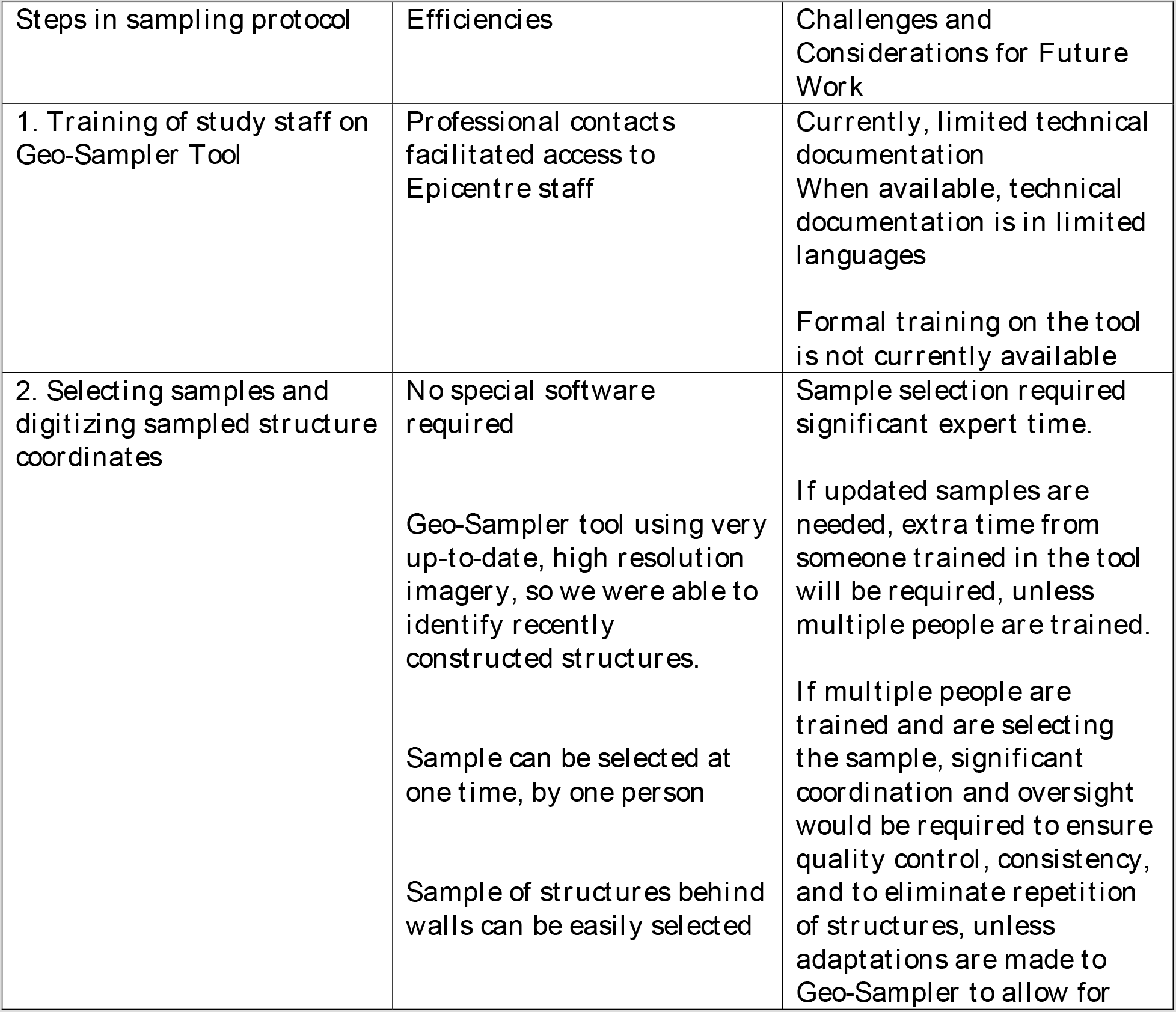
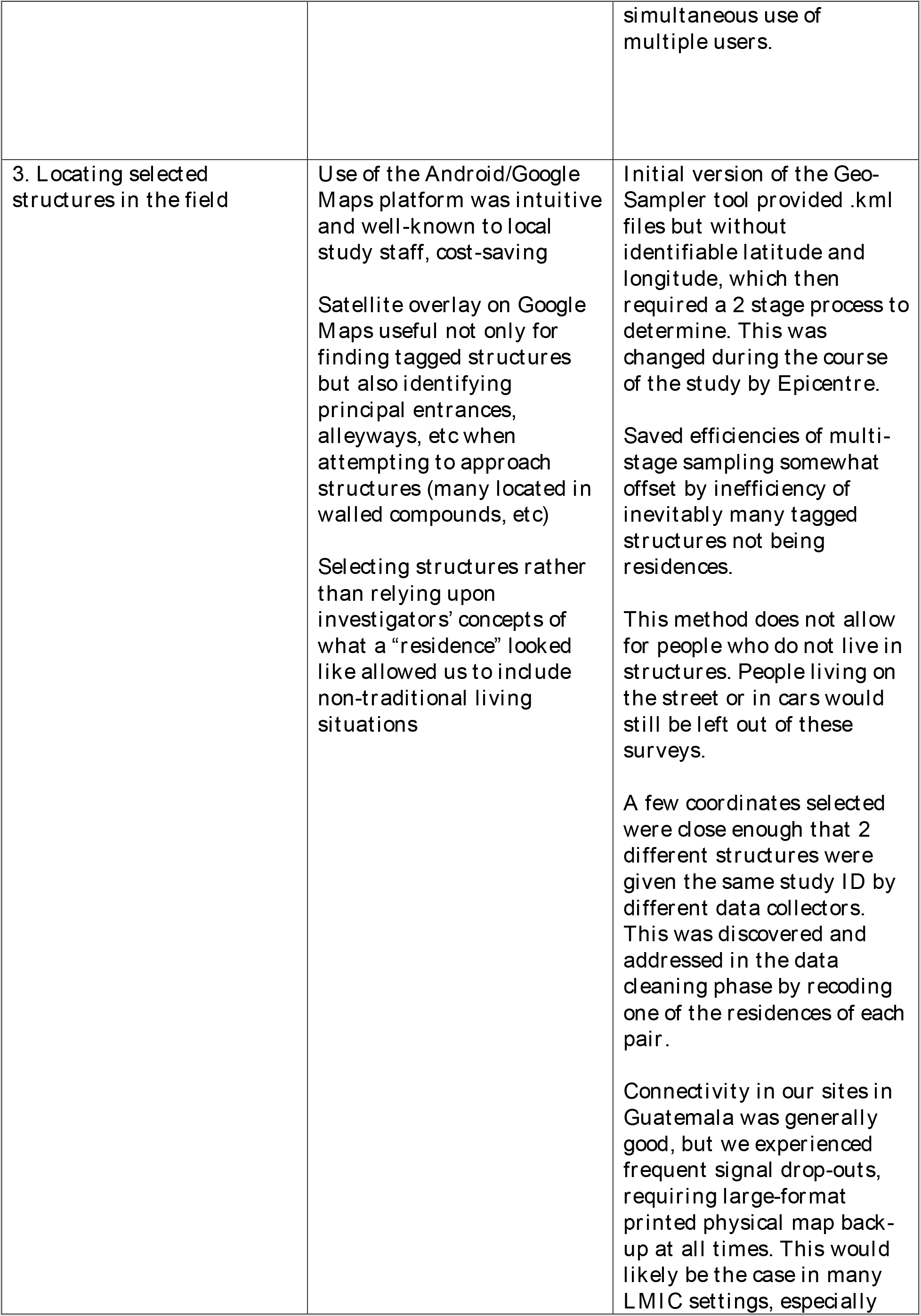
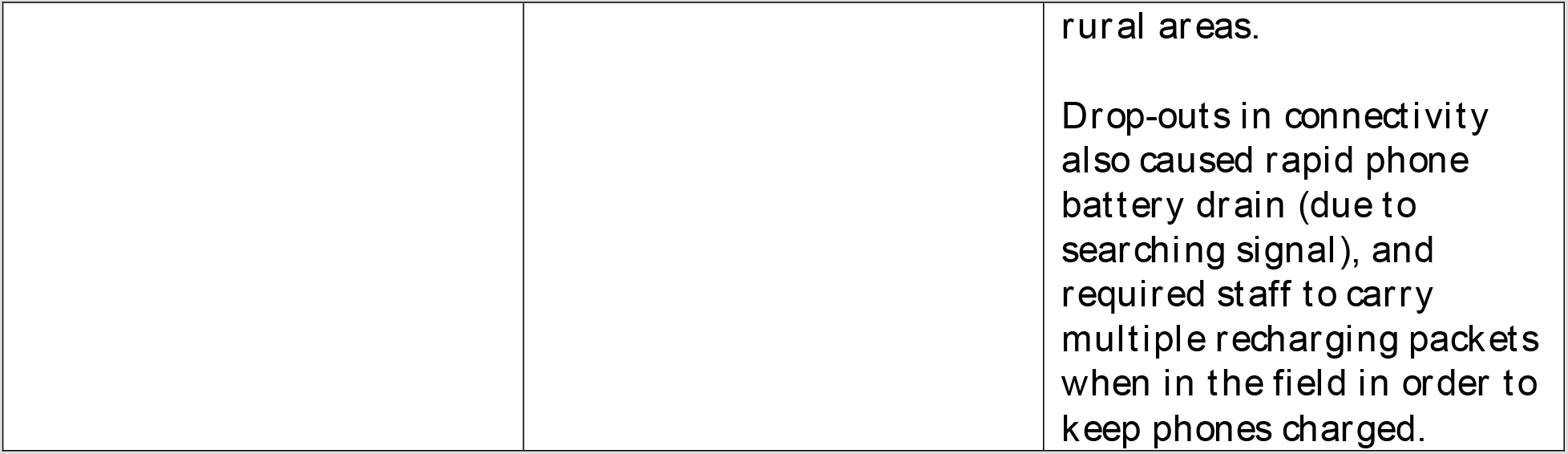
Efficiencies and Challenges of using Geo-Sampler tool and protocols for population-level data collection in Guatemala.

| Steps in sampling protocol | Efficiencies | Challenges and Considerations for Future Work |
| --- | --- | --- |
| 1. Training of study staff on Geo-Sampler Tool | Professional contacts facilitated access to Epicentre staff | Currently, limited technical documentation<br>When available, technical documentation is in limited languages<br>Formal training on the tool is not currently available |
| 2. Selecting samples and digitizing sampled structure coordinates | No special software required<br>Geo-Sampler tool using very up-to-date, high resolution imagery, so we were able to identify recently constructed structures.<br>Sample can be selected at one time, by one person<br>Sample of structures behind walls can be easily selected | Sample selection required significant expert time.<br>If updated samples are needed, extra time from someone trained in the tool will be required, unless multiple people are trained.<br>If multiple people are trained and are selecting the sample, significant coordination and oversight would be required to ensure quality control, consistency, and to eliminate repetition of structures, unless adaptations are made to Geo-Sampler to allow for |
|  |  | simultaneous use of multiple users. |
| 3. Locating selected structures in the field | Use of the Android/Google Maps platform was intuitive and well-known to local study staff, cost-saving<br>Satellite overlay on Google Maps useful not only for finding tagged structures but also identifying principal entrances, alleyways, etc when attempting to approach structures (many located in walled compounds, etc)<br>Selecting structures rather than relying upon investigators' concepts of what a "residence" looked like allowed us to include non-traditional living situations | Initial version of the Geo-Sampler tool provided .kml files but without identifiable latitude and longitude, which then required a 2 stage process to determine. This was changed during the course of the study by Epicentre.<br>Saved efficiencies of multi-stage sampling somewhat offset by inefficiency of inevitably many tagged structures not being residences.<br>This method does not allow for people who do not live in structures. People living on the street or in cars would still be left out of these surveys.<br>A few coordinates selected were close enough that 2 different structures were given the same study ID by different data collectors. This was discovered and addressed in the data cleaning phase by recoding one of the residences of each pair.<br>Connectivity in our sites in Guatemala was generally good, but we experienced frequent signal drop-outs, requiring large-format printed physical map back-up at all times. This would likely be the case in many LMIC settings, especially |
|  |  | rural areas.<br>Drop-outs in connectivity also caused rapid phone battery drain (due to searching signal), and required staff to carry multiple recharging packets when in the field in order to keep phones charged. |

**Table 3:** Data collected on each sampled structure.

| Query | Possible Responses |
| --- | --- |
| Is the selected structure a residence?<sup>1</sup> | Yes/No |
| How many associated structures are in use by the household? | Number of structures and type<sup>2</sup> |
| Does more than one household live in structure? | Number of households in residence |
| Recruitment outcomes | Number of households approached<br>Number of households with a contact<br>Number of households with at least one member recruited (“hh enrolled”)<br>Number of eligible adults in household and whether each individual was recruited or declined participation |

<sup>1</sup>Including structures of multiple use (stores, churches, etc) as long as also a residence.
<sup>2</sup>For example, selected structure might be the primary residence for the household, but there may also be a separate kitchen structure and garage structure for the same household

The data collection field team initially consisted of two community nurses with LPN-equivalent degrees and one supervisor with masters-level in nursing. While this level of training was necessary for our study to collect biologic samples and conduct interviews, advanced degrees are not necessary to use Geo-Sampler outputs to identify households. Only familiarity with smartphone technology, map literacy, and Google Maps is required. A two day training and pilot exercise was sufficient to allow the data collectors to get started. Time between map generation and first data collection in the field was two days.

Data cleaning was conducted throughout the study by two people at a time; one of the coinvestigators (ACM) and the project coordinator (serially, ED LS, ET) with support from the PI (PR). All data cleaners and analysts had a Masters or PhD. Skill required for data cleaning include familiarity with excel, Redcap and Stata programming. Calculation of sample weights after data cleaning took approximately three days. Calculation of sample weights involved sophisticated demography skills for devising the formulae, and strong MS excel skills to program the spreadsheets. Application of the sampling and response weights to generate prevalence estimates required advanced Stata programming skills, but once the programs were created, they could be reused swiftly. Investigators on the team(ACM, DRT, PR) have strong excel skills, statistical programming software skills (Stata 15), and/or advanced population health and demography skills.

## Discussion

We have identified several advantages of this method over the more traditional methods of sampling frame enumeration in this area of Guatemala. The main advantages are those of time, cost and statistical efficiency.

The traditional enumeration method of household census or sending a mapping crew ahead of time to knock on doors is time-consuming and expensive. DHS suggests estimating two months for this phase in their field manuals(22), and another two months between household enumeration and data collection. In a smaller survey of 1600 households in rural Madagascar, enumeration required 20 days with 9 field teams, and another 24 days between enumeration and the initiation of data collection(23). Our survey required a total of 1 week’s hours (38.5) of full-time work (across six non-consecutive weeks) by one person to generate the maps plus one week of training and two days from beginning of sample selection to beginning of field time. In this area of Guatemala, many households are located behind privacy or security walls and residents may not admit enumeration crew to their homes, leaving the crew to simply guess at the number of households behind the walls. This area of Guatemala is not heavily forested, so satellite imagery gives an excellent representation of the structures available for sampling. Additionally, Geo-Sampler’s use of high-resolution satellite imagery captured within six months of the initiation of the field work provided a much more up-to-date source of population data typically used to select cluster surveys.

Furthermore, surveys which digitized all structures from satellite imagery as a sample frame required intimate knowledge of the area including typical structures and living patterns to be able to produce a reasonable map of residential buildings in the area. The digitization process is quite time-consuming and relies on the two-step process of first enumeration followed by selection. The Geo-Sampler method does not require digitizing all structures, and allows for simultaneous enumeration and selection, at a very rapid pace. Because each point in the polygon has an equal chance of being selected, and the user has the ability to select only points with a structure contained within the buffer as part of the sample, statistical correction for clustering is not required. In this lower-resource context, the absence of both 1)a recent national census and the corresponding updated sampling frame and 2) an updated accurate gridded population dataset, Geo-Sampler allowed us to select a simple random sample of households from a random sample of structures, rather than a multi-stage clustered sample, which allowed us to reduce our sample size and conduct the study with a reduced budget.

This method does have some possible limitations when implemented. The areas of Guatemala in which the study was conducted vary greatly in terms of environment, climate, and wealth. However, they are similar in their levels of arboreal cover and that most of the buildings are single family houses. Although a small proportion of the population in each site reside in apartment buildings or other multi-family dwellings, the vast majority of families reside in single-family houses. These tools and protocols are well-suited to this scenario, but may be less applicable to densely populated urban settings with multiple living units per structure.

Despite the overall successful implementation of the protocols, our study team did face a few challenges in the use of the protocols during the field work. The study coordinator used a mobile device to identify a selected structure, visited it first to determine if it was actually a household, and if so, enrolled any eligible participants. The study nurses then followed up at a pre-arranged time to conduct study activities. On a few occasions, the randomly selected coordinates were close enough that two different structures were given the same study ID by different data collectors. This was discovered during the cleaning process and one of the households was reassigned to a different number. However, this was a time-consuming puzzle. Connectivity was also an issue; the data collectors experienced frequent signal drop-outs, and used a lot of battery life in searching for connectivity. This issue will probably be applicable to other lower-resource settings as well. We addressed this by providing large-format printed maps as a back-up. Harnessing the knowledge locally hired staff have of their neighborhoods is also important, and including names of neighborhoods was useful to support the ability of nurses to easily find houses. Although data collectors had extensive experience with smart phones, some study staff were less comfortable using both paper maps and the phone-based Google Maps. Despite these challenges, nurse data collectors have been successfully able to locate and identify the selected households for inclusion into the study, replacing with others as necessary.

Although these methods provide a step forward in including non-traditional living arrangements (people residing permanently or temporarily in businesses, religious institutions or other structures), they do not capture some of the most marginalized and vulnerable people in the population—the unhoused, street dwellers or people living in vehicles. Some of the structures selected were in areas determined to be too dangerous to the safety of data collectors to enroll. Thus, there remains a group of community residents whose health indicators, perspectives and risks are invisible and undocumented. Further research on ways to include these most vulnerable groups in population studies is needed.

### Conclusion

In combination with our design protocols, the Epicentre Geo-Sampler tool provided a feasible, rapid and lower-cost alternative to select a representative population sample for a prevalence survey in our setting.

## Data Availability

Datasets created for this manuscript are not publicly available because they are coordinates of specific structures and residences of study participants; these data will not be made public for confidentiality reasons. However, a deidentified replication dataset from the entire study (Chronic Kidney Disease of unknown etiology) will be made available at the Principal Investigator's Harvard University's Dataverse following publication of final study results.

## List of Abbreviations

CKD: Chronic Kidney disease
CKDu: Chronic Kidney Disease of unknown etiology
DHS: Demographic and Health Surveys
EPI: Expanded Programme on Immunization
GPS: Geographic positioning system
KDIGO: Kidney Disease: Improving Global Outcomes
LMIC: Low- and middle-income countries
LPN: Licensed practical nurse
MICS: Multiple Indicator Cluster Surveys
NIH: National Institutes of Health

## DECLARATIONS

### Ethics approval and consent to participate

The study was reviewed and approved by the Partners Health Care, Maya Health Alliance, and the Instituto de Nutricion de Centro América y Panama’s respective Institutional Review Board.

### Consent for publication

All authors have reviewed the manuscript and agree to its publication.

### Availability of data and materials

Datasets created for this manuscript are not publically available because they are coordinates of specific structures and residences of study participants; these data will not be made public for confidentiality reasons. However, a deidentified replication dataset from the entire CKDu study will be made available at the Principal Investigator’s Harvard University’s Dataverse following publication of final study results.

### Authors’ Contributions

PR: Principal Investigator of the CKDu study; designed study, contributed to data acquisition, co-wrote first and all drafts of the manuscript

ACM: designed study, conducted sample selection, all data analysis and interpretation, wrote first and all drafts of manuscript

AB: helped test the Geo-Sampler tool, interpreted results, contributed to multiple drafts of manuscript, reviewed and approved final draft

ED: assisted in design of field study, contributed to data collection and cleaning, reviewed and approved final draft of manuscript

LS: contributed to data collection and cleaning, contributed to multiple drafts of manuscript, reviewed and approved final draft

ET: led final data cleaning, contributed to analysis, reviewed and approved final draft of manuscript

FG: was involved in design and application of Geo-Sampler tool, and reviewed the final draft of the manuscript

CMM: Co-PI of the CKDu study, assisted with interpretation of results, reviewed and approved final draft of manuscript

DRT: contributed to design of study, designed analysis and mentored ACM on conduct of analysis, contributed to interpretation of result, co-wrote first and all drafts of manuscript

## Acknowledgements

The authors gratefully acknowledge the hard work of the data collection teams, specifically Irma Yolanda Raquec Teleguario, Glenda Gómez Hernández, and Mérida Isabel Coj Sajvin, Héctor Gomez Hernández, Alvaro Danilo Sir López and staff at Wuqu’ Kawok. We acknowledge the support of the INCAP laboratories, and Dr. Joaquin Barnoya for productive conversations. We gratefully acknowledge the work of Serge Balandine from Epicentre on design of Geo-Sampler and Epicentre for their cooperation in allowing us to use this tool. We especially thank the study participants, without whom this would not be possible.

